# Safety of the single-dose Ad26.CoV2.S vaccine among healthcare workers in the phase 3b Sisonke study in South Africa

**DOI:** 10.1101/2021.12.20.21267967

**Authors:** Simbarashe Takuva, Azwidhwi Takalani, Ishen Seocharan, Nonhlanhla Yende-Zuma, Tarylee Reddy, Imke Engelbrecht, Mark Faesen, Kentse Khuto, Carmen Whyte, Veronique Bailey, Valentina Trivella, Jonathan Peter, Jessica Opie, Vernon Louw, Pradeep Rowji, Barry Jacobson, Pamela Groenewald, Rob E. Dorrington, Ria Laubscher, Debbie Bradshaw, Harry Moultrie, Lara Fairall, Ian Sanne, Linda Gail-Bekker, Glenda Gray, Ameena Goga, Nigel Garrett, the Sisonke study team

## Abstract

**Background:** The Sisonke open-label phase 3b implementation study aimed to assess the safety and effectiveness of the Janssen Ad26.CoV2.S vaccine among health care workers (HCWs) in South Africa. Here, we present the safety data.

**Methods:** We monitored adverse events (AEs) at vaccination sites, through self-reporting triggered by text messages after vaccination, health care provider reports and by active case finding. The frequency and incidence rate of non-serious and serious AEs were evaluated from day of first vaccination (17 February 2021) until 28 days after the final vaccination (15 June 2021). COVID-19 breakthrough infections, hospitalisations and deaths were ascertained via linkage of the electronic vaccination register with existing national databases.

**Findings:** Of 477,234 participants, 10,279 (2.2%) reported AEs, of which 139 (1.4%) were serious. Women reported more AEs than men (2.3% vs. 1.6%). AE reports decreased with increasing age (3.2% for 18–30, 2.1% for 31-45, 1.8% for 46-55 and 1.5% in >55-year-olds). Participants with previous COVID-19 infection reported slightly more AEs (2.6% vs. 2.1%). The commonest reactogenicity events were headache and body aches, followed by injection site pain and fever, and most occurred within 48 hours of vaccination. Two cases of Thrombosis with Thrombocytopenia Syndrome and four cases of Guillain-Barre Syndrome were reported post-vaccination. Serious AEs and AEs of special interest including vascular and nervous system events, immune system disorders and deaths occurred at lower than the expected population rates.

**Interpretation:** The single-dose Ad26.CoV2.S vaccine had an acceptable safety profile supporting the continued use of this vaccine in our setting.

**Funding:** Funding was provided by the National Treasury of South Africa, the National Department of Health, Solidarity Response Fund NPC, The Michael & Susan Dell Foundation, The Elma Vaccines and Immunization Foundation - Grant number 21-V0001, and the Bill & Melinda Gates Foundation – grant number INV-030342.

## BACKGROUND

South Africa is among the most affected countries globally by COVID-19, with over 266,000 excess natural deaths occurring between May 2020 and October 2021 (approximately 448 per 100,000 individuals).(1) The single-dose Ad26.COV2.S vaccine showed efficacy in preventing symptomatic and severe COVID-19 disease in the ENSEMBLE study(2) including in South Africa, where initially the beta and then the delta variants were the predominant circulating strains.(3) Here, an estimated 1,300 health care workers (HCWs) have died from COVID-19 as of September 2021.(4) The Sisonke study, a phase 3b open-label single arm implementation study of the single-dose Ad26.COV2.S vaccine was conducted as an emergency intervention to protect HCWs in the face of an anticipated third COVID-19 wave, at a time when no vaccines were available through the national rollout.

The Ad26.COV2.S vaccine is compatible with standard vaccine storage and distribution channels and is therefore a practical vaccine for low- and middle-income countries or remote populations.(5) To date, approximately 30 million persons in the United States (US) and the European Union have received the Ad26.COV2.S vaccine.(6) Vaccine adverse event (AE) surveillance systems demonstrate that billions of people have safely received COVID-19 vaccines.(7) Adverse events following COVID-19 vaccination are generally mild, and include local reactions, such as injection site pain, redness, swelling, and systemic reactions, like fever, headache, fatigue, nausea, vomiting, diarrhea.(8,9)

As reported by the US Center for Disease Control, severe or potentially life-threatening AEs are rare, and after 12.6 million doses of the Ad26.CoV2.S vaccine, 38 cases of Thrombosis with Thrombocytopenia Syndrome (TTS) and 98 cases of Guillain-Barré Syndrome (GBS) were reported, while after 141 million second mRNA vaccine doses, 497 cases of myocarditis were reported.(10) Following the precautionary pause instituted by the Food and Drug Administration (FDA) in April 2021, the SA Health Products Regulatory Authority recommended a similar 2-week pause for the Sisonke study.(11) The study recommenced with additional safeguards including screening and monitoring of participants at high risk of thrombosis and implementing measures to safely manage participants with TTS. Participant information sheets and informed consent forms were updated to include the newly identified AEs. Identification of such rare events illustrated that continued evaluation of the safety profile of vaccines post licensure is crucial to accurately characterize safety and to identify very rare adverse events that may not be reported in clinical trials.

The Sisonke study enrolled almost half a million HCWs, providing an opportunity to further evaluate the safety of the Ad26.COV.S2 vaccine in an expanded population.

## METHODS

### Study participants

The Sisonke study is a multi-centre, open-label, single-arm phase 3b implementation study among HCWs (>18 years) in South Africa, which is conducted in collaboration with the National Department of Health (ClinicalTrials.gov number NCT04838795; Pan-African Clinical Trials registry number PACTR202102855526180). All eligible HCWs, who registered on the national Electronic Vaccination Data System (EVDS) and provided electronic consent for the study, were eligible for enrolment. Pregnant and breastfeeding women were excluded due to a lack of sufficient safety data at that time. A total of 477,234 HCWs received the Ad26.COV.S2 vaccine between 17 February 2021 and 17 May 2021.

The institutional health research ethics committees of participating clinical research sites approved the study, which was overseen by the South African Health Products Regulatory Authority (Ref: 20200465).

### Vaccination procedures

Participants received appointments for vaccination through the EVDS or were invited via employer lists. Vaccinations were conducted in collaboration with the National Department of Health public or private vaccination centres across all nine South African provinces and overseen by Good Clinical Practice-trained personnel linked to one of the ENSEMBLE trial research sites. Participants received a single intramuscular injection of Ad26.COV2.S at a dose of 5×10^10^ virus particles and were observed for AEs for 15 minutes post-vaccination, and for 30 minutes, if they had a previous history of allergic reactions to vaccinations.

### Adverse event reporting

Adverse events were reported into the study database via multiple streams using a hybrid surveillance system that combined passive with active reporting.(12) Firstly, we designed an electronic case report form (eCRF). After vaccination every participant received a text message with COVID-19 infection prevention measures, that also listed common signs and symptoms of reactogenicity and provided an AE reporting web link, which allowed participants access to the form for AE reporting. Second, health care providers were able to complete paper-based CRFs that were available at healthcare and vaccination facilities, which were then submitted to the Sisonke safety desk and captured in the AE database. Third, the study team set up a safety desk call centre staffed by pharmacovigilance nurses, pharmacists and safety physicians to assess and advise on AE reports. Contact details were advertised on the vaccination cards, shared on social media and included in the text messages. Finally, spontaneous case reports via unsolicited communication by HCWs were captured and verified by safety desk staff.

In addition, we actively linked EVDS data via national identification numbers with national patient-level disease databases, COVID-19 case notifications and the national population registry to identify vaccinees with COVID-19 infections, COVID-19-related hospitalisations and deaths. COVID-19 is a notifiable medical condition in South Africa and tests conducted across laboratories are reposited in the National Health Laboratory Service data system, which was used to identify seropositive Sisonke participants via active linkage. A death notification form must be submitted to the Department of Home Affairs to obtain a death certificate. Therefore, in addition to case reports and active tracing, mortality was ascertained via linkage with the national population registry. After identification of deaths, the safety staff contacted next of kin, primary health care providers and solicited medical records to ascertain causes of death.

### Safety monitoring

Adverse event reports were processed daily and screened for serious AEs (SAE), which met International Conference on Harmonization criteria and AEs of special interest (AESI) per Brighton Collaboration list (9). Seven days after reporting an AE, participants received a follow-up text message with a link to the eCRF. Participants reporting worsening and non-resolving symptoms were followed up by safety desk staff. After the FDA lifted the cautionary 2-week pause in vaccinations, two additional follow-up text messages were sent to all participants seven and 14 days after vaccination. The texts highlighted signs/symptoms associated with TTS and provided a link to the eCRF. Safety staff made attempts to obtain medical records and supporting information from health care providers for all reported SAEs.

The protocol safety review team comprising principal investigators, safety physicians and subject matter experts (haematologists, neurologist, allergy expert, infectious disease specialists) provided oversight by weekly safety data review. An independent safety monitoring committee provided additional safety oversight.

### Statistical analysis

For descriptive statistics, counts and proportions were used for categorical variables, and medians and interquartile ranges for quantitative variables. Participants reporting and not reporting AEs were compared by baseline characteristics. SAEs were summarized by MedDRA System Organ Class and AE Preferred Term. For selected SAEs, disproportionality analysis was conducted: the observed (O) number of reported cases was compared to the expected (E) number based on background incidence rates and the O/E ratio with 95% confidence interval was calculated. Available background incidence rates were used including a medically insured population in South Africa (pulmonary embolism and deep venous thrombosis), Tanzanian population-based cohort study (neurological events such as stroke) and European population databases.(10, 15–18) Person-time was accrued from the date of vaccination until death or dataset closure on 15 June 2021 (28 days after the last vaccination). The incidence rates per 100,000 person-years were calculated using a Poisson model with person-years as an offset. Deaths were excluded from the SAE analysis and examined as a separate entity. COVID-related deaths were excluded in this report and published in a separate effectiveness report. All statistical analyses were conducted using STATA version 14 (STATA Corp., College Station, TX, USA).

## RESULTS

### Participants

The Sisonke study enrolled and vaccinated 477,234 participants from all nine South African provinces between 17 February 2021 and 17 May 2021. The majority were women (74.9%), the median age was 42 years (IQR 33-51) and 16.3% were older than 55 years. Previous COVID-19 infection was self-reported by 14.5% of participants. The most prevalent comorbidities were hypertension (15.6%), HIV infection (8.3%) and diabetes mellitus (5.9%).

### Adverse events reported by baseline characteristics

A total of 10,279 AEs were reported, of which 138 (1.4%) were classified as serious. Most AE reports (81%) were electronic self-reports. Women reported more AEs than men (2.3% versus 1.6%; p< 0.001), AE reporting decreased with increasing age (3.2% for 18 to 30-year-olds versus 1.5% in ≥ 55-year-olds; p<0.001), and participants with previous COVID-19 infection reported more AEs (2.6% versus 2.1%; p<0.001). Persons living with HIV (1.2% versus 2.2%; p<0.001) or previous tuberculosis (0.8% versus 2.2%; p=0.043) reported less AEs than those without, while more AEs were reported by participants with chronic lung disease compared to those without (4.7% versus 2.1%; p<0.001). Proportions reporting AEs among those with other comorbidities were similar. (**Table 1)**.

**Table 1:** Demographic and clinical characteristics of Ad26.CoV2.S vaccine recipients in the Sisonke Study.

| Characteristics | Total |  | Not reporting AEs |  | Reporting AEs |  | P-value |
| --- | --- | --- | --- | --- | --- | --- | --- |
|  | N | % | N | % | N | % |  |
| **Sex** |  |  |  |  |  |  | <0.001 |
| Female | 357481 | 74.9 | 349092 | 97.7 | 8389 | 2.3 |  |
| Male | 119753 | 25.1 | 117863 | 98.4 | 1890 | 1.6 |  |
| **Age (Median, IQR)** | 42 | 33-51 | 42 | 33-51 | 38 | 30-48 | <0.001 |
| **Age category (years)** |  |  |  |  |  |  | <0.001 |
| 18-30 | 85486 | 17.9 | 82789 | 96.8 | 2697 | 3.2 |  |
| 31-45 | 209376 | 43.9 | 204892 | 97.9 | 4484 | 2.1 |  |
| 46-55 | 104078 | 21.8 | 102176 | 98.2 | 1902 | 1.8 |  |
| >55 | 78162 | 16.4 | 76967 | 98.5 | 1195 | 1.5 |  |
| **Previous COVID-19** |  |  |  |  |  |  | <0.001 |
| No | 408085 | 85.5 | 399607 | 97.9 | 8478 | 2.1 |  |
| Yes | 69149 | 14.5 | 67348 | 97.4 | 1801 | 2.6 |  |
| **Comorbidities** |  |  |  |  |  |  |  |
| Hypertension |  |  |  |  |  |  | <0.001 |
| No | 402853 | 84.4 | 394007 | 97.8 | 8846 | 2.2 |  |
| Yes | 74381 | 15.6 | 72948 | 98.1 | 1433 | 1.9 |  |
| Diabetes |  |  |  |  |  |  | 0.003 |
| No | 449171 | 94.1 | 439427 | 97.8 | 9744 | 2.2 |  |
| Yes | 28063 | 5.9 | 27528 | 98.1 | 535 | 1.9 |  |
| HIV |  |  |  |  |  |  | <0.001 |
| No | 437848 | 91.7 | 428048 | 97.8 | 9800 | 2.2 |  |
| Yes | 39386 | 8.3 | 38907 | 98.8 | 479 | 1.2 |  |
| Cancer |  |  |  |  |  |  | 0.528 |
| No | 47587 | 99.7 | 465617 | 97.8 | 10253 | 2.2 |  |
| Yes | 1364 | 0.3 | 1338 | 98.1 | 26 | 1.9 |  |
| Previous Tuberculosis |  |  |  |  |  |  | 0.047 |
| No | 476756 | 99.9 | 466481 | 97.8 | 10275 | 2.2 |  |
| Yes | 478 | 0.1 | 474 | 99.2 | 4 | 0.8 |  |
| Heart disease |  |  |  |  |  |  | 0.232 |
| No | 473804 | 99.3 | 463609 | 97.8 | 10195 | 2.2 |  |
| Yes | 3430 | 0.7 | 3346 | 97.6 | 84 | 2.4 |  |
| Chronic lung disease |  |  |  |  |  |  | <0.001 |
| No | 475501 | 99.6 | 465304 | 97.9 | 10197 | 2.1 |  |
| Yes | 1733 | 0.4 | 1651 | 95.3 | 82 | 4.7 |  |

Most reported AEs (n=9,021, 81%) were reactogenicity events; the commonest were headaches and body aches (including arthralgia, myalgia and fatigue) which occurred within the first seven days of vaccination, followed by mild injection site pain and fever (**Figure 1**). These events occurred predominately on the day of vaccination, reducing in frequency by day two. Self-reported severity was mild to moderate in 67% (n=7,157), i.e. the event did not result in loss of ability to perform usual social and functional activities, while 32% (n=3,375) reported being unable to perform usual activities and 2% (n=213) reported that they needed to visit the emergency room or were hospitalised. Follow up at day seven post vaccination indicated that 92% of participants reporting AEs had either completely recovered or were recovering. The remaining 8% of participants were contacted by the safety team and, if required, referred for care. One in five (19%) AEs were not consistent classified as reactogenicity events (**Supplementary Table 1)**.

**Figure 1.**
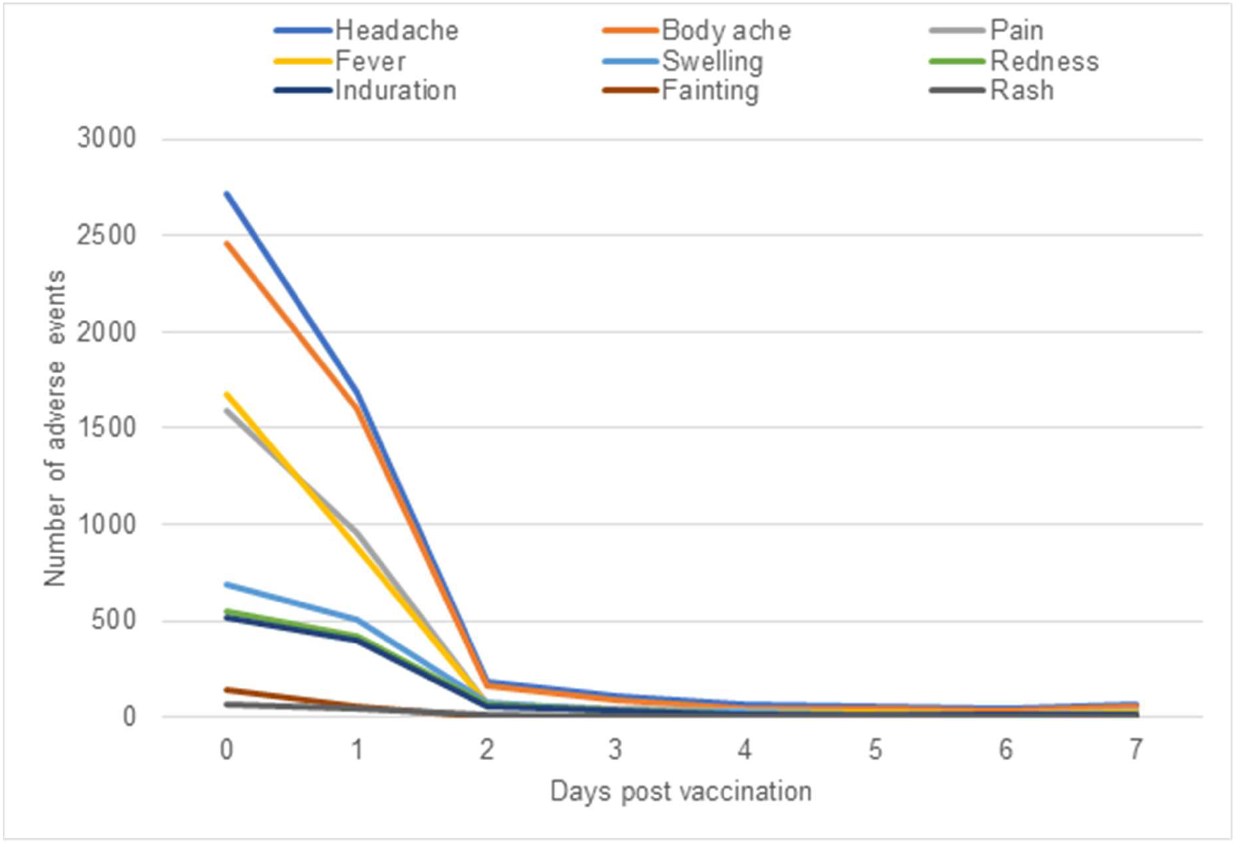
Commonly occurring adverse events in the first seven days post vaccination.

### Serious adverse events

A total of 138 SAEs (excluding deaths) were reported by 136 participants (median age 42 years, IQR 35-51) with 114 (82.0%) reported by women and 25 (18.0%) by men. Most SAEs (115; 82.7%) occurred within 28 days of vaccination with a median time to onset (all SAEs) of 5 days (IQR 1-17) and for SAEs occurring with 28 days of vaccination, 1 day (IQR 0-9). Vascular (n=37; 39.1/ 100,000 person-years) and nervous system disorders (n=31; 31.66/ 100,000 person-years), infections and infestations (n=19; 24.3/ 100,000 person-years) and immune system disorders (n=24; 20.1/ 100,000 person-years) were the commonest reported SAE categories (**Table 2**). SAE outcomes were: 48 (34.8%) recovered, 36 (26.1%) recovering, 45 (32.6%) ongoing and 9 (6.5%) deceased.

**Table 2:**
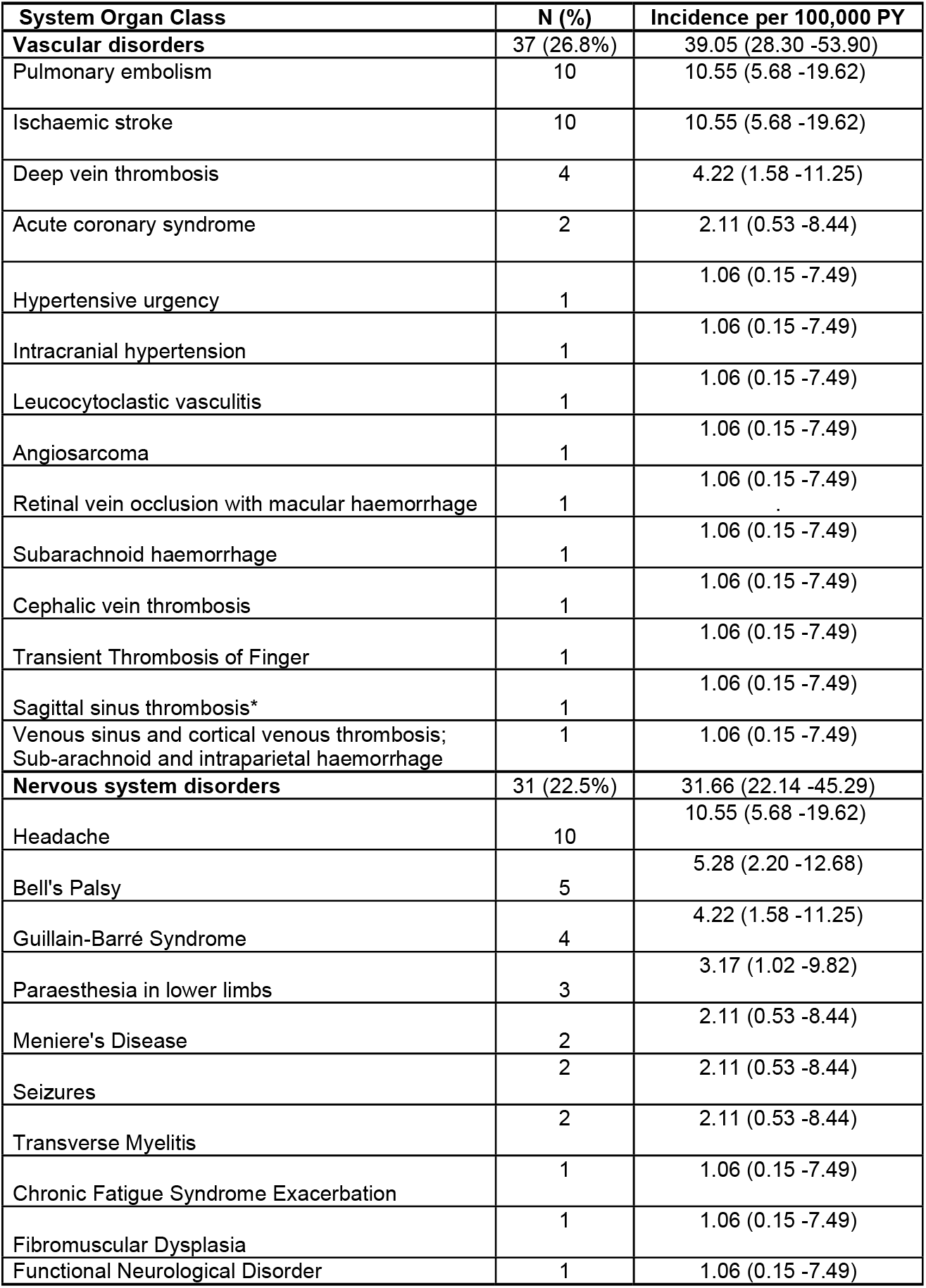
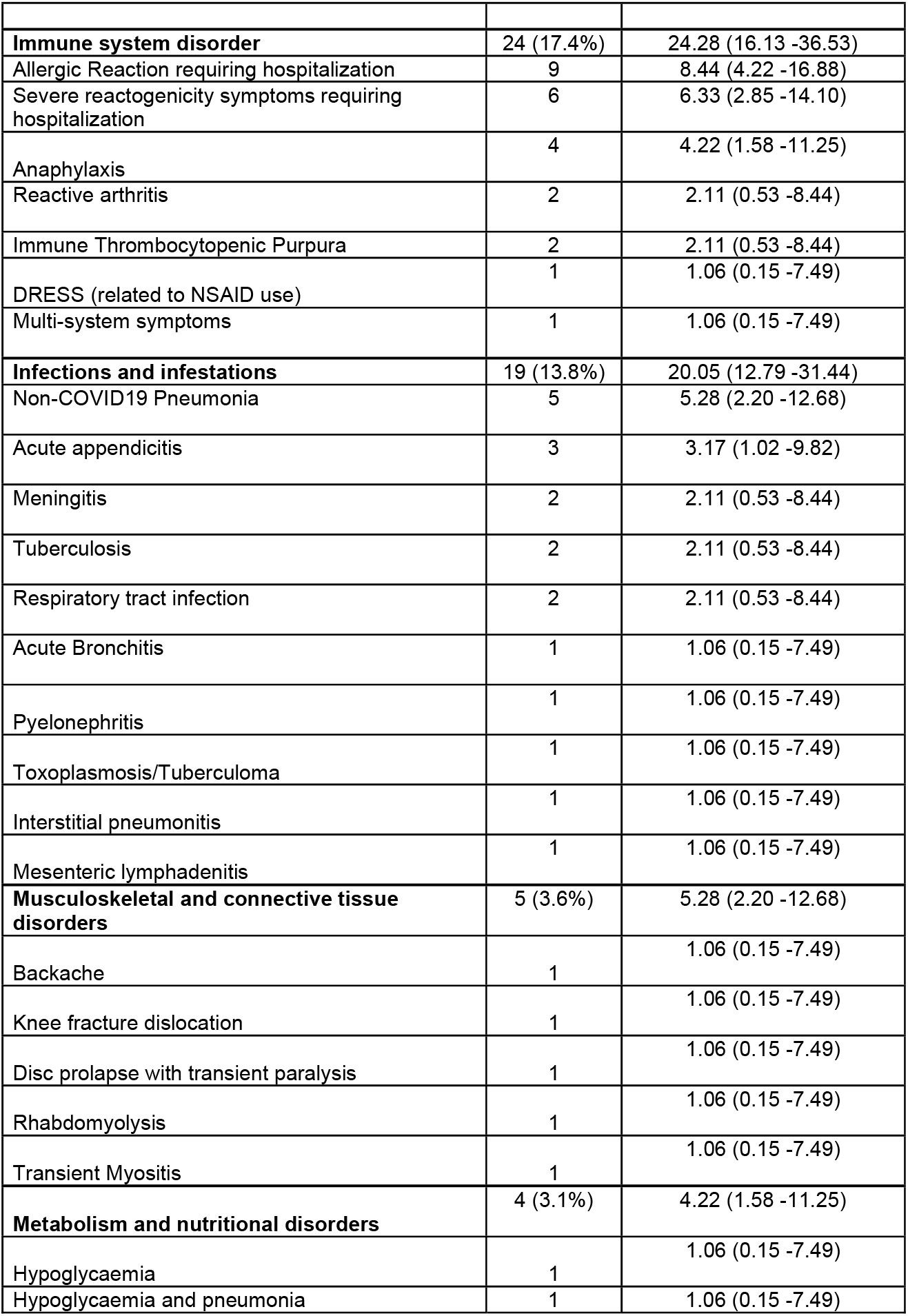
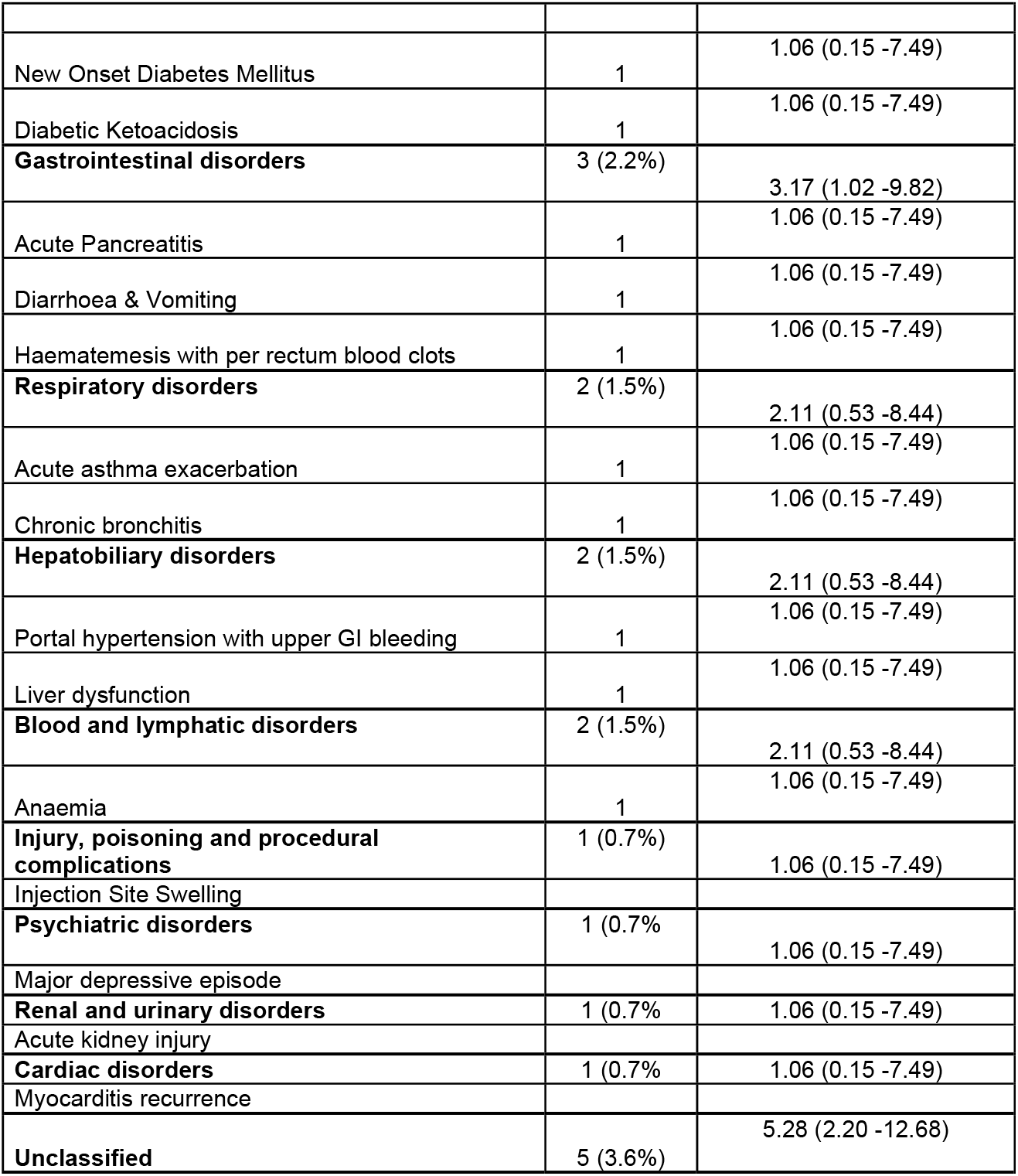
Serious adverse events by Medical Dictionary for Regulatory Activities (MEDRA) System Organ Class and Preferred Adverse Event term (N=138)

| **System Organ Class** | **N (%)** | **Incidence per 100,000 PY** |
| --- | --- | --- |
| **Vascular disorders** | 37 (26.8%) | 39.05 (28.30 -53.90) |
| Pulmonary embolism | 10 | 10.55 (5.68 -19.62) |
| Ischaemic stroke | 10 | 10.55 (5.68 -19.62) |
| Deep vein thrombosis | 4 | 4.22 (1.58 -11.25) |
| Acute coronary syndrome | 2 | 2.11 (0.53 -8.44) |
| Hypertensive urgency | 1 | 1.06 (0.15 -7.49) |
| Intracranial hypertension | 1 | 1.06 (0.15 -7.49) |
| Leucocytoclastic vasculitis | 1 | 1.06 (0.15 -7.49) |
| Angiosarcoma | 1 | 1.06 (0.15 -7.49) |
| Retinal vein occlusion with macular haemorrhage | 1 | 1.06 (0.15 -7.49) |
| Subarachnoid haemorrhage | 1 | 1.06 (0.15 -7.49) |
| Cephalic vein thrombosis | 1 | 1.06 (0.15 -7.49) |
| Transient Thrombosis of Finger | 1 | 1.06 (0.15 -7.49) |
| Sagittal sinus thrombosis* | 1 | 1.06 (0.15 -7.49) |
| Venous sinus and cortical venous thrombosis;<br>Sub-arachnoid and intraparietal haemorrhage | 1 | 1.06 (0.15 -7.49) |
| **Nervous system disorders** | 31 (22.5%) | 31.66 (22.14 -45.29) |
| Headache | 10 | 10.55 (5.68 -19.62) |
| Bell's Palsy | 5 | 5.28 (2.20 -12.68) |
| Guillain-Barré Syndrome | 4 | 4.22 (1.58 -11.25) |
| Paraesthesia in lower limbs | 3 | 3.17 (1.02 -9.82) |
| Meniere's Disease | 2 | 2.11 (0.53 -8.44) |
| Seizures | 2 | 2.11 (0.53 -8.44) |
| Transverse Myelitis | 2 | 2.11 (0.53 -8.44) |
| Chronic Fatigue Syndrome Exacerbation | 1 | 1.06 (0.15 -7.49) |
| Fibromuscular Dysplasia | 1 | 1.06 (0.15 -7.49) |
| Functional Neurological Disorder | 1 | 1.06 (0.15 -7.49) |
| **Immune system disorder** | 24 (17.4%) | 24.28 (16.13 -36.53) |
| Allergic Reaction requiring hospitalization | 9 | 8.44 (4.22 -16.88) |
| Severe reactogenicity symptoms requiring hospitalization | 6 | 6.33 (2.85 -14.10) |
| Anaphylaxis | 4 | 4.22 (1.58 -11.25) |
| Reactive arthritis | 2 | 2.11 (0.53 -8.44) |
| Immune Thrombocytopenic Purpura | 2 | 2.11 (0.53 -8.44) |
| DRESS (related to NSAID use) | 1 | 1.06 (0.15 -7.49) |
| Multi-system symptoms | 1 | 1.06 (0.15 -7.49) |
| **Infections and infestations** | 19 (13.8%) | 20.05 (12.79 -31.44) |
| Non-COVID19 Pneumonia | 5 | 5.28 (2.20 -12.68) |
| Acute appendicitis | 3 | 3.17 (1.02 -9.82) |
| Meningitis | 2 | 2.11 (0.53 -8.44) |
| Tuberculosis | 2 | 2.11 (0.53 -8.44) |
| Respiratory tract infection | 2 | 2.11 (0.53 -8.44) |
| Acute Bronchitis | 1 | 1.06 (0.15 -7.49) |
| Pyelonephritis | 1 | 1.06 (0.15 -7.49) |
| Toxoplasmosis/Tuberculoma | 1 | 1.06 (0.15 -7.49) |
| Interstitial pneumonitis | 1 | 1.06 (0.15 -7.49) |
| Mesenteric lymphadenitis | 1 | 1.06 (0.15 -7.49) |
| **Musculoskeletal and connective tissue disorders** | 5 (3.6%) | 5.28 (2.20 -12.68) |
| Backache | 1 | 1.06 (0.15 -7.49) |
| Knee fracture dislocation | 1 | 1.06 (0.15 -7.49) |
| Disc prolapse with transient paralysis | 1 | 1.06 (0.15 -7.49) |
| Rhabdomyolysis | 1 | 1.06 (0.15 -7.49) |
| Transient Myositis | 1 | 1.06 (0.15 -7.49) |
| **Metabolism and nutritional disorders** | 4 (3.1%) | 4.22 (1.58 -11.25) |
| Hypoglycaemia | 1 | 1.06 (0.15 -7.49) |
| Hypoglycaemia and pneumonia | 1 | 1.06 (0.15 -7.49) |
| New Onset Diabetes Mellitus | 1 | 1.06 (0.15 -7.49) |
| Diabetic Ketoacidosis | 1 | 1.06 (0.15 -7.49) |
| **Gastrointestinal disorders** | 3 (2.2%) | 3.17 (1.02 -9.82) |
| Acute Pancreatitis | 1 | 1.06 (0.15 -7.49) |
| Diarrhoea & Vomiting | 1 | 1.06 (0.15 -7.49) |
| Haematemesis with per rectum blood clots | 1 | 1.06 (0.15 -7.49) |
| **Respiratory disorders** | 2 (1.5%) | 2.11 (0.53 -8.44) |
| Acute asthma exacerbation | 1 | 1.06 (0.15 -7.49) |
| Chronic bronchitis | 1 | 1.06 (0.15 -7.49) |
| **Hepatobiliary disorders** | 2 (1.5%) | 2.11 (0.53 -8.44) |
| Portal hypertension with upper GI bleeding | 1 | 1.06 (0.15 -7.49) |
| Liver dysfunction | 1 | 1.06 (0.15 -7.49) |
| **Blood and lymphatic disorders** | 2 (1.5%) | 2.11 (0.53 -8.44) |
| Anaemia | 1 | 1.06 (0.15 -7.49) |
| **Injury, poisoning and procedural complications** | 1 (0.7%) | 1.06 (0.15 -7.49) |
| Injection Site Swelling |  |  |
| **Psychiatric disorders** | 1 (0.7%) | 1.06 (0.15 -7.49) |
| Major depressive episode |  |  |
| **Renal and urinary disorders** | 1 (0.7%) | 1.06 (0.15 -7.49) |
| Acute kidney injury |  |  |
| **Cardiac disorders** | 1 (0.7%) | 1.06 (0.15 -7.49) |
| Myocarditis recurrence |  |  |
| **Unclassified** | 5 (3.6%) | 5.28 (2.20 -12.68) |

The commonest vascular disorders were pulmonary emboli (n=10, 10.6 per 100,000 person years, 95%CI 5.7-19.6) and ischaemic strokes (n=10, 10.6 per 100,000 person years, 95%CI 5.7-19.6) followed by deep vein thrombosis (n=4, 4.2 per 100,000 person-years, 95%CI 1.6-11.3). Three participants had both pulmonary embolism and deep vein thrombosis. There were two cases classified as TTS. The first case was a woman in the 45-50 year age group presenting with pulmonary embolism, thrombocytopenia and positive anti-platelet factor 4 antibodies nine days after vaccination. She had a history of injectable contraceptive use, underlying chronic respiratory, neurological condition and was being investigated for an autoimmune disorder. The second case was a woman in the 25-30 year age group who was admitted to hospital unconscious after experiencing a severe headache, restlessness and confusion from 33 days after vaccination. A CT brain scan with venogram was in keeping with superior sagittal sinus thrombosis. Anti-platelet factor 4 antibodies assay were negative, and she had marginally low platelets. She was a current smoker but had no other significant medical history. Both participants recovered.

Most events affecting the nervous system were complaints of headaches or migraines resulting in hospital admissions (n=8). Five cases of Bell’s palsy (5.3 per 100,000 person-years, 95%CI 2.2-12.7) were reported between one and 42 days after vaccination: Two men (40-45 year age group) developed GBS about two weeks after vaccination and two women (50-55 year age group) developed GBS 16 and 17 days after vaccination (4.2 per 100,000 person-years, 95%CI 1.6-11.3). All participants are recovering. Four cases were adjudicated as anaphylaxis(13) by two physicians using the Brighton Collaboration and National Institutes for Allergy and Infectious Diseases case definition; with cases needing to meet both definitions to be considered confirmed cases.(17) All anaphylaxis cases had previous occurrence of drug or vaccine-associated anaphylaxis and recovered fully. There was one case of myocarditis in a woman with previous myocarditis, which had settled prior to vaccination. She is receiving care.

**Table 3** summarizes the disproportionality analysis that compares the occurred versus expected incidence ratio for AEs of concern. Out of the AEs examined, TTS and GBS occurred at rates greater than the baseline comparison population (O/E ratio 2.4; 95%CI 0.3-8.7) and O/E ratio 5.1; 95%CI 1.4-13.0 respectively). For the other AEs, namely, ischaemic stroke, pulmonary embolism (non-TTS), deep vein thrombosis, acute coronary syndrome, Bell’s palsy, transverse myelitis, seizures and myocarditis, the O/E ratio was <1. **Supplementary Table 2** describes the disproportionate analysis for SAEs occurring within 28 days of vaccination and **Supplementary Figure 1** illustrates the frequency of SAE reporting from day of vaccination. As expected, there was a drop off in events when we analysis was restricted to 28 days post-vaccination, but however there was not a significant change in the observed vs. expected ratios of SAEs.

**Table 3:** Observed versus expected analysis of selected serious adverse events.

| Adverse event | Observed count | Observed incidence rate per 100,000 PY | Expected count | Expected incidence rate per 100,000 PY | O/E ratio (95% CI) |
| --- | --- | --- | --- | --- | --- |
| **Vascular disorders** |  |  |  |  |  |
| Ischaemic stroke | 10 | 10.55 (5.68 -19.62) | 102.89 | 108.60 (14) | 0.10 (0.05 – 0.18) |
| Pulmonary embolism | 10 | 10.55 (5.68 -19.62) | 21.09 | 22.26 (15) | 0.47 (0.23 – 0.87) |
| Deep vein thrombosis | 4 | 4.22 (1.58 -11.25) | 30.50 | 32.19 (15) | 0.13 (0.04 – 0.34) |
| Acute coronary syndrome | 2 | 2.11 (0.53 -8.44) | 214.12 | 226.00 (16) | 0.01 (0.00 – 0.03) |
| Thrombotic thrombocytopenic syndrome | 2 | 2.11 (0.53 -8.44) | 0.83 | 0.88 (17) | 2.40 (0.29-8.66) |
| **Neurological disorders** |  |  |  |  |  |
| Bell's palsy | 5 | 5.28 (2.20 - 12.68) | 21.32 | 22.50 (19) | 0.23 (0.08-0.55) |
| Guillain-Barré syndrome | 4 | 4.22 (1.58 -11.25) | 0.79 | 0.83 (20) | 5.09 (1.39-13.02) |
| Transverse myelitis | 2 | 2.11 (0.53 -8.44) | 28.14 | 29.70 (18) | 0.08 (0.01 – 0.27) |
| Seizure | 2 | 2.11 (0.53 -8.44) | 69.45 | 73.30 | 0.03 (0.00-0.10) |
| **Cardiac disorders** |  |  |  |  |  |
| Myocarditis | 1 | 1.06 (0.15 -7.49) | 20.84 | 22.00 (21) | 0.05 (0.00 – 0.27) |

The OE analysis compares the observed and expected numbers of cases. This may be expressed as the ratio
The rates are for adults (male/females) and are not stratified by age – group.
Where a range is given in the literature on incidence, the mid-point was used.

A total of 157 non-COVID-19 related deaths (167 per 100,000 person-years) were identified via the active linkage system with the national population registry. Of these deaths, 67/157 (42.7%) causes of death were adjudicated and ascertainment continues for the remainder. 38% (n=60) were male, median age was 48 years (IQR 40-57), 42 (26.8%) were reported as non-natural causes and comorbidities reported were as follows: hypertension (n=48, 30.6%), diabetes (n=32, 20.4%), HIV (n=20, 12.7%), heart disease (n=9, 5.7%), cancer (n=1, 1.7%) and 57% (n=90) had at least one comorbidity. Adjudicated causes of death included metastatic cancer (n=18), HIV/AIDS-related deaths (n=15), motor vehicle accidents (n=11), homicide (n=7), pulmonary embolism (n=5), myocardial infarction (n=4), cerebrovascular accident/stroke (n=3), non-COVID-19 pneumonia (n=3), intracerebral bleeds (n=3), bleeding peptic ulcer/upper GIT bleed (n=3), suicide (n=2), status epilepticus (n=2). Other causes are shown in **Supplementary Table 3**.

Nineteen deaths occurred within 28 days after vaccination. Causes were motor vehicle accident (3), upper gastrointestinal tract bleeding (3), homicide (2), HIV/AIDS related deaths (2), one each of pulmonary embolism, metastatic pancreatic cancer, drowning, dilated cardiomyopathy, renal failure, myelodysplastic syndrome, status epilepticus, suicide and a death post aortic valve and bypass surgery. A woman (25-30 year age group), with a history of hypertension post-delivery, presented 20 days after vaccination to her physician with jaundice and anuria. She then developed confusion, renal failure and haemolysis requiring dialysis and fresh frozen plasma transfusion. She demised after transfer to an intensive care unit. Investigations were in keeping the Thrombotic Thrombocytopenic Purpura (TTP). She was HIV negative, and no other triggers could be identified. Assessment of the event using the World Health Organization Causality Assessment Tool (18), classified this as an indeterminate temporal relationship with insufficient evidence for attribution to the vaccine. In the absence of a clear alternative cause, the safety team deemed it plausible that vaccine could have exacerbated this event in a patient with a predisposition to TTP.

Finally, we compared age-standardized mortality rates in Sisonke with projected background population mortality rates in South Africa as per the 2018 *Medical Research Council Rapid Mortality Surveillance Report* and pre-COVID-19 local employee group life assurance data for a similar age-structured working population.(19) The mortality rate in Sisonke was similar to the working population mortality data with similar ages, and well below that of the overall population mortality rate (**Figure 2**).

**Figure 2.**
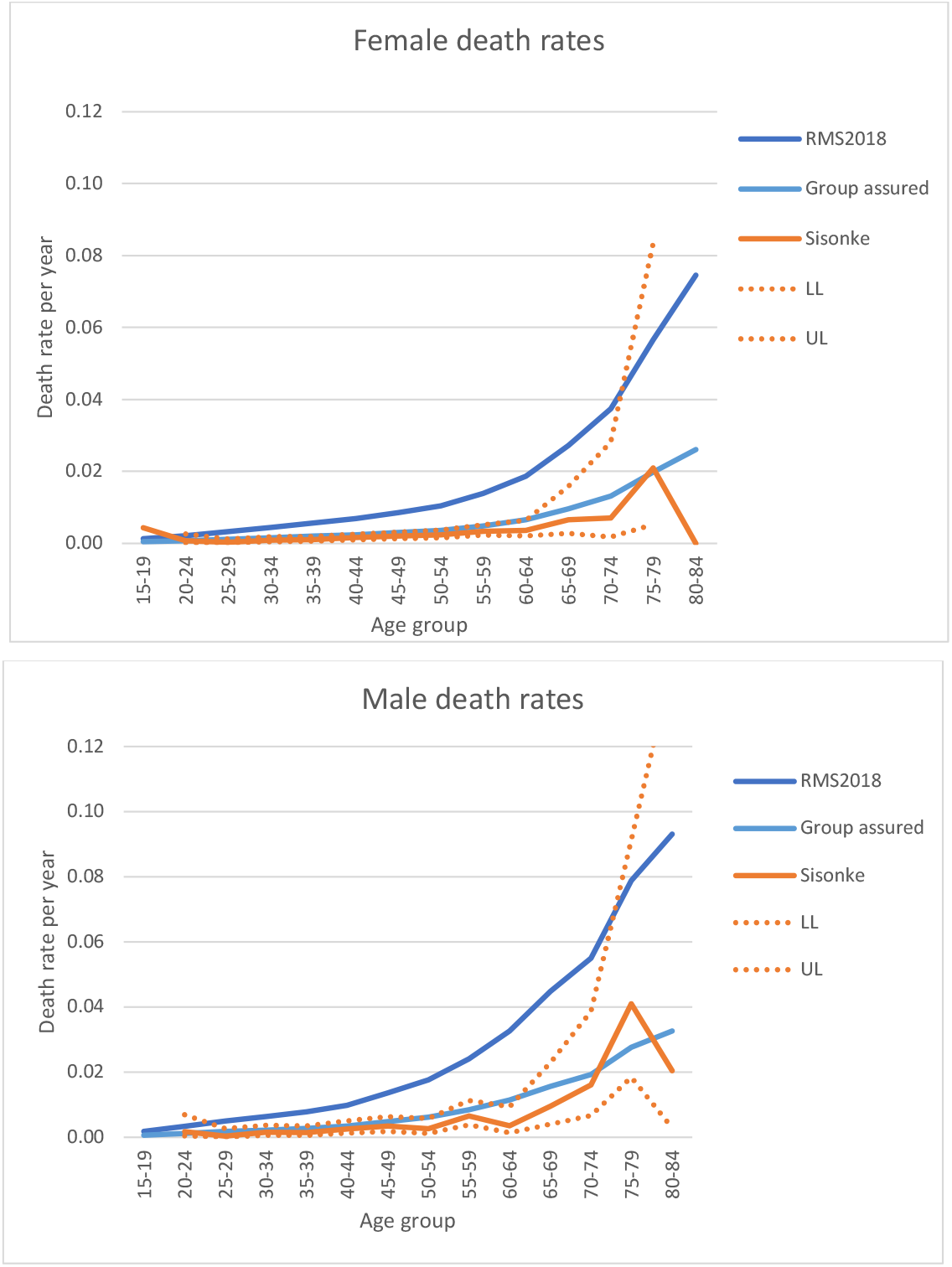
Age-standardized mortality rates by sex in the Sisonke study compared to 2018 South Africa mortality rates and working population mortality rates. Age-standardized mortality rates for projected background population in SA as per the 2018 Medical Research Council Rapid Mortality Surveillance Report and pre-COVID-19 local employee group life assurance data for a similar age-structured working population.

## DISCUSSION

The Sisonke study, a large single arm, open-label phase 3b implementation study aimed to assess the safety and effectiveness of the single-dose Ad26.CoV2.S vaccine among almost half a million HCWs in South Africa. With over 10,000 AE reports, this was the largest safety analysis of the Ad26.CoV2.S vaccine from a low- and middle-income country. As observed in phase 3 trials, similar patterns of AEs were found and were mostly expected reactogenicity signs and symptoms. Furthermore, SAEs were rare and occurred below expected rates. However, we did observe very rare events of TTS and GBS in this study. Nevertheless, overall, this study provides additional real-world evidence that the vaccine is safe and well tolerated, supporting its continued use in this setting.

Adverse events were more often reported by women than men. While this may illustrate a stronger immune response seen in females compared to males(20–22), behavioural factors such as reduced reporting among men may have played a role, but were not measured. The prevalence of reported AEs reduced with increasing age. A number of studies show that vaccine-related AEs and reactogenicity are less prevalent in older people due to waning of innate immune defence mechanisms, lower systemic levels of IL-6, IL-10, C-reactive protein and lower neutralising antibody titres after vaccination as compared to younger individuals.(23–25) Individuals reporting previous COVID-19 infection seemed to have more AE reporting rates. Some studies suggest that there is increased immunogenicity in the setting of past infection leading to higher antibody titres(26,27) and therefore higher reactogenicity rates.

Thrombosis with Thrombocytopenia Syndrome and GBS occurred at very low rates in this study, however the disproportionality analysis showed a higher event rate than expected in the population. After 12.6 million doses of the Ad26.CoV.2 vaccine had been administered in the US, 38 confirmed TTS cases were reported and 98 cases of GBS.(10) Based on this data, estimates illustrate a clear advantage of vaccination despite these rare AE occurrences. For example, in the 30-49 year age group among women, for every 6-7 cases of GBS or 8-10 cases of TTS, 10,100 COVID-19 cases, 900 hospitalizations, 140 ICU admissions and 20 deaths were prevented.(10)

While the risk-benefit balance clearly favours vaccination, this study highlights the importance of ongoing safety monitoring in population-based vaccination programmes to enable continued risk-benefit assessment. The Sisonke study shows that additional surveillance, heightened awareness, and development of protocols for the management of potential clinical complications after vaccination, help identify and manage possible cases early and appropriately. For example, the two cases of TTS were successfully managed with the support of the protocol safety review team and both participants recovered. It is crucial that such cases are identified promptly to enable successful management. Local clinical recommendations for management of TTS were developed and implemented.(28)

Overall, the Sisonke study did not identify excess deaths in the vaccinated population compared to the general population. Mortality rates were comparable to a similar adult working population from 2018. This report illustrates the importance of accurate national mortality surveillance, especially in settings where vaccine hesitancy is driven by non-scientific and inaccurate reports in communities and through social media. The Sisonke results are also a strong reminder that South Africa faces a large burden of disease.(29) While cancer was the commonest cause of death during the study period, which highlights the urgent need for specialist oncology services, it is concerning that among HCWs advanced HIV and tuberculosis remain common causes of death. It shows that despite gains in access to HIV testing and treatment, HIV and TB care require further improvement. Local data highlight that the COVID-19 epidemic heavily impacted HIV testing and treatment initiations.(30) Motor vehicle accidents and homicides were also common causes of death, a reflection of the injury and trauma burden in South Africa. One death was related to TTP, which has previously been reported post Ad26.COV2-S vaccination(31) and warrants further evaluation in other studies.

The Sisonke study had several limitations. Firstly, the surveillance system was primarily passive relying on self-reporting, thus some AEs may have gone unreported. It is likely that the system was better suited to detect SAEs than milder AEs, which participants may have ignored rather than reported. As active contact with participants was up to two-weeks post-vaccination, it is probable that SAEs other than deaths and COVID-19 events were more likely to be reported during this period leading to underestimation of SAEs which occurred. The active linkage of the unique identifier in EVDS with the deaths on the national population registry and COVID-19 laboratory system ensured identification of nearly all possible deaths and COVID-19 events in the study. Additionally, considering the large number of participants in the study, not all self-reported AEs could be verified and only SAEs and AEs of medical concern were investigated further. Interpretation of the disproportionality analysis should be cautious given the uncertainties in both the observed and expected event rates, variable follow-up time, non-South African reference data for some groups, and potential differences in age-sex distributions between Sisonke and reference data. However, while disproportionality analysis in the context of safety signal detection is mainly exploratory, it has the utility to identify potentially important associations between AE and vaccine. In this study, the analysis confirmed current reports on safety risk of the Ad26.CoV2.S vaccine with the TTS and GBS.(10) Lastly, It is also important to note that without a placebo group, open label single arm studies are subject to measurement bias with the potential of overreporting of AEs and hence some caution in interpreting safety data. No other safety concerns were found in this study.(10, 11)

In conclusion, this study affirms that the single dose Ad26.COV2.S vaccine is safe and well tolerated when administered to adults in South Africa. Few SAEs were observed and they were successfully managed with prompt identification. The Sisonke study underscores the value of setting up robust pharmacovigilance systems for prompt identification, evaluation and reporting of AEs to enable continued assessment of the risk-benefit profiles of COVID-19 vaccines. This has the potential to improve public confidence in vaccine safety and reduce vaccine hesitancy.

## Data Availability

All data produced in the present study are available upon reasonable request to the authors

## ACKNOWLEDGEMENTS

We acknowledge the South African Medical Research Council for sponsorship and oversight, Janssen Vaccines and Prevention for the supply and transport of the study product to South Africa, Right to Care for supporting the safety monitoring and reporting infrastructure and the National Department of Health.

We thank all the health care workers that participated in this study, the investigators and staff members at the vaccination centers and clinical research sites, the protocol team, protocol safety review team and members of the independent data and safety monitoring committee (Sipho Dlamini, Yunus Moosa, Francesca Conradie, Chris Breyer, Jeremy Nel).

The Sisonke study was funded by the South African Medical Research Council and funds were received from National Treasury through the South African National Department of Health and from the Solidarity Fund, the ELMA Vaccines and Immunization Foundation and the Michael & Susan Dell Foundation.

## TABLE AND FIGURES

**Supplementary Table 1:** Twenty commonest non-reactogenicity adverse events reports.

| **Adverse event** | **Frequency** |
| --- | --- |
| Nausea and /or Vomiting | 592 |
| Dizziness | 450 |
| Flu-like symptoms | 344 |
| Chills and rigors | 336 |
| Diarrhoea | 294 |
| Chest pain | 262 |
| Allergy like symptoms | 250 |
| Abdominal symptoms with or without diarrhoea | 190 |
| Cough | 169 |
| Numbness and Paraesthesia | 168 |
| Tachycardia | 114 |
| Allergy like symptoms | 102 |
| Itchiness | 93 |
| Tight chest or shortness of breath | 61 |
| Loss of taste or smell | 41 |
| Bruised on injection site | 35 |
| Lymphadenopathy | 33 |
| Insomnia | 31 |
| Elevated BP | 23 |
| Asthma exacerbation or bronchial spasm | 13 |
| Blister/s | 13 |
| Breast pain/swelling | 13 |

The 20 commonest non-reactogenicity AEs are shown in Supplemental Table 1

**Supplementary Table 2:** Observed versus expected analysis of selected serious adverse events within 28 days of vaccination. Total person-years: 37 915.

| **Adverse event** | **Observed count** | **Observed incidence rate per 100,000 PY** | **Expected count** | **Expected incidence rate per 100,000 PY** | **O/E ratio (95% CI)** |
| --- | --- | --- | --- | --- | --- |
| **Vascular disorders** |  |  |  |  |  |
| Ischaemic stroke | 8 | 21.10 (10.55 - 42.19) | 41.18 | 108.60 (14) | 0.19 (0.08 – 0.38) |
| Pulmonary embolism** | 6 | 15.82 (7.11 -35.22) | 8.44 | 22.26 (15) | 0.71 (0.26 – 1.54) |
| Deep vein thrombosis | 4 | 10.55 (3.96 -28.11) | 12.20 | 32.19 (15) | 0.33 (0.08 – 0.84) |
| Acute coronary syndrome | 1 | 2.64 (0.37 -18.72) | 85.69 | 226.00 (16) | 0.01 (0.00 – 0.07) |
| Thrombotic thrombocytopenic syndrome | 1 | 2.64 (0.37 -18.72) | 0.33 | 0.88 (17) | 3.0 (0.08-16.70) |
| **Neurological disorders** |  |  |  |  |  |
| Bell's palsy | 2 | 5.27 (1.32 -21.09) | 8.53 | 22.5 (19) | 0.23 (0.03-0.85) |
| Guillain-Barré syndrome | 4 | 10.55 (3.96 -28.11) | 0.31 | 0.83 (20) | 12.71 (3.46-32.54) |
| Transverse myelitis | 0 | - | 28.14 | 29.7 (18) | 0.08 (0.01 – 0.27) |
| Seizure | 1 | 2.64 (0.37 -18.72) | 27.79 | 73.3 | 0.04 (0.00-0.20) |
| **Cardiac disorders** |  |  |  |  |  |
| Myocarditis | 1 | 2.64 (0.37 -18.72) | 8.34 | 22 (21) | 0.12 (0.003 – 0.67) |

**Supplementary Figure 1.**
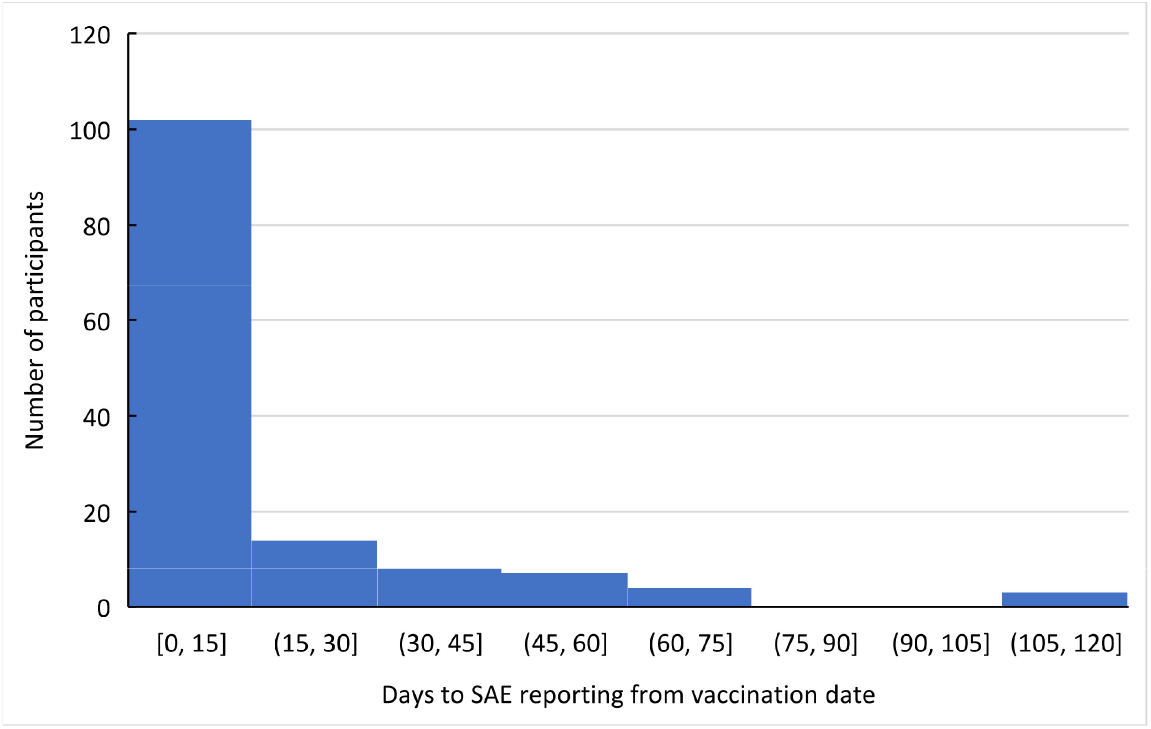
Frequency of SAE reports from time of vaccination.

**Supplementary Table 3:** Mortality in the Sisonke study.

|  | **Overall (n=157)** | **Cause known (n=66)** | **Unknown (n=91)** |
| --- | --- | --- | --- |
| Male sex, n (%) | 60 (38.0%) | 28 (41.8%) | 32 (35.2%) |
| Age (years), median (IQR) | 48 (40 – 47) | 48 (41 – 58) | 48 (40 – 57) |
| Time to death, median (IQR) |  |  |  |
| ≤ 28 days, n (%) | 53 (33.7%) | 19 (35.8%) | 34 (64.2%) |
| 28 days to ≤ 3 months | 88 (56.1%) | 42 (47.7%) | 46 (52.3%) |
| > 3 months | 16 (10.2%) | 5 (31.2%) | 11 (31.2%) |
| Cause listed as non-natural, n (%) | 42 (26.6%) | 15 (22.4%) | 27 (29.7%) |
| Comorbidities, n (%) |  |  |  |
| Hypertension | 48 (30.4%) | 21 (31.3%) | 27 (29.7%) |
| Diabetes | 32 (20.3%) | 13 (19.4%) | 19 (20.9%) |
| HIV | 20 (12.7%) | 12 (17.9%) | 8 (8.8%) |
| Total number comorbidities | 90 (57.0%) | 44 (65.7%) | 46 (50.6%) |

## REFERENCES

1. Bradshaw, D., Laubscher, R., Dorrington, R., Groenewald, P. & Moultrie T. Report on Weekly Deaths in South Africa: 3 May 2020 -16 Oct 2021. Cape Town; 2021.

2. Sadoff J, Gray G, Vandebosch A, Cárdenas V, Shukarev G, Grinsztejn B, et al. Safety and Efficacy of Single-Dose Ad26.COV2.S Vaccine against Covid-19. N Engl J Med. 2021 Jun;384(23):2187–201.

3. Tegally H, Wilkinson E, Giovanetti M, Iranzadeh A, Fonseca V, Giandhari J, et al. Detection of a SARS-CoV-2 variant of concern in South Africa. Nature [Internet]. 2021;592(7854):438–43. Available from: http://dx.doi.org/10.1038/s41586-021-03402-9

4. Division S. COVID-19 Sentinel Hospital Surveillance Weekly Update on Hospitalized HCWs. 2021;(March 2020):1–10.

5. Janssen Biotech I. Factsheet: Emergency Use Authorization (EUA) of the Janssen COVID-19 vaccine to prevent coronavirus disease, 2019 [Internet]. Vol. 2019. Horsham, PA; 2019. Available from: https://www.fda.gov/media/146305/download

6. University of Oxford. Covid vaccine doses by manufacturer [Internet]. Our World in Data. 2021. Available from: https://ourworldindata.org/grapher/covid-vaccine-doses-by-manufacturer?country=~USA

7. Shimabukuro TT, Nguyen M, Martin D, DeStefano F. Safety monitoring in the Vaccine Adverse Event Reporting System (VAERS). Vaccine. 2015 Aug;33(36):4398–405.

8. Biotech J. Vaccines and Related Biological Products Advisory Committee Meeting FDA Briefing Document Janssen Ad26. COV2. S Vaccine for the Prevention of COVID-19 Sponsor : 2021;

9. Barbara L, Miriam S. Safety Platform for Emergency vACcines. Priority List of Adverse Events of Special Interest: COVID-19. 2020;(December):V2.0.

10. Rosenblum H. COVID-19 Vaccines in Adults : Benefit-Risk Discussion Current COVID-19 vaccine policy. 2021; SAHPRA.

11. SAHPRA Statement – Update on Sisonke Phase 3B Implementation Study [Internet]. 2021. p. 2021. Available from: https://www.sahpra.org.za/wp-content/uploads/2021/04/SAHPRA-Statement-on-pausing-of-the-Sisonke-study-19-April-.pdf

12. Takuva S, Takalani A, Garrett N, Goga A, Peter J, Louw V, et al. Thromboembolic Events in the South African Ad26.COV2.S Vaccine Study. The New England journal of medicine. 2021.

13. Law B. Safety Platform for Emergency vACcines Guide for 1st Tier AESI Acute Myelitis. 2020;(February):0–32. Available from: https://brightoncollaboration.us/wp-content/uploads/2021/03/SPEAC_D2.5.2.1_Anaphylaxis-Case-Definition-Companion-Guide_V1.0-12070-1.pdf

14. Pitsavos C, Panagiotakos DB, Antonoulas A, Zombolos S, Kogias Y, Mantas Y, et al. Epidemiology of acute coronary syndromes in a Mediterranean country; aims, design and baseline characteristics of the Greek study of acute coronary syndromes (GREECS). BMC Public Health [Internet]. 2005;5(1):23. Available from: https://doi.org/10.1186/1471-2458-5-23

15. Discovery Health Medical Scheme. Deep venous thrombosis and pulmonary embolism rates, South Africa.

16. Duarte-salles T, Fernandez-bertolin S, Aragón M, Reyes C, Martinez-E. Background rates of five thrombosis with thrombocytopenia syndromes of special interest for COVID-19 vaccine safety surveillance: incidence between 2017 and 2019 and patient profiles from 20.6 million people in six European countries. medRxiv [Internet]. 2021; Available from: https://www.medrxiv.org/content/10.1101/2021.05.12.21257083v1.full.pdf

17. Manivannan V, Decker WW, Stead LG, Li JTC, Campbell RL. Visual representation of national institute of allergy and infectious disease and food allergy and anaphylaxis network criteria for anaphylaxis. Int J Emerg Med. 2009;2(1):3–5.

18. WHO. Causality Assessment Of An Adverse Event Following Immunization [Internet]. 2019. 1–62 p. Available from: https://apps.who.int/iris/bitstream/handle/10665/259959/9789241513654-eng.pdf)

19. Bradshaw D, Dorrington R. Rapid Mortality Surveillance Report 2011. 2012.

20. Klein SL, Marriott I, Fish EN. Sex-based differences in immune function and responses to vaccination. Trans R Soc Trop Med Hyg. 2015 Jan;109(1):9–15.

21. Flanagan KL, Fink AL, Plebanski M, Klein SL. Sex and Gender Differences in the Outcomes of Vaccination over the Life Course. Annu Rev Cell Dev Biol. 2017 Oct;33:577–99.

22. Markle JG, Fish EN. SeXX matters in immunity. Trends Immunol. 2014 Mar;35(3):97– 104.

23. Ramasamy MN, Minassian AM, Ewer KJ, Flaxman AL, Folegatti PM, Owens DR, et al. Safety and immunogenicity of ChAdOx1 nCoV-19 vaccine administered in a prime-boost regimen in young and old adults (COV002): a single-blind, randomised, controlled, phase 2/3 trial. Lancet (London, England). 2021 Dec;396(10267):1979–93.

24. Sadoff J, Le Gars M, Shukarev G, Heerwegh D, Truyers C, de Groot AM, et al. Interim Results of a Phase 1-2a Trial of Ad26.COV2.S Covid-19 Vaccine. N Engl J Med. 2021 May;384(19):1824–35.

25. Walsh EE, Frenck RWJ, Falsey AR, Kitchin N, Absalon J, Gurtman A, et al. Safety and Immunogenicity of Two RNA-Based Covid-19 Vaccine Candidates. N Engl J Med. 2020 Dec;383(25):2439–50.

26. Manisty C, Otter AD, Treibel TA, McKnight Á, Altmann DM, Brooks T, et al. Antibody response to first BNT162b2 dose in previously SARS-CoV-2-infected individuals. Vol. 397, Lancet (London, England). 2021. p. 1057–8.

27. Wise J. Covid-19: People who have had infection might only need one dose of mRNA vaccine. BMJ. 2021 Feb;372:n308.

28. Jacobson BF, Schapkaitz E, Mer M, Louw S, Haas S, Buller HR, et al. Recommendations for the diagnosis and management of vaccine-induced immune thrombotic thrombocytopenia. South African Med J. 2021;111(6):535–7.

29. Bradshaw D, Nannan N, Pillay-Van Wyk V, Laubscher R, Groenewald P, Dorrington RE. Burden of disease in South Africa: Protracted transitions driven by social pathologies. South African Med J. 2019;109(11b):69–76.

30. Dorward J, Khubone T, Gate K, Ngobese H, Sookrajh Y, Mkhize S, et al. The impact of the COVID-19 lockdown on HIV care in 65 South African primary care clinics: an interrupted time series analysis. Lancet HIV [Internet]. 2021;8(3):e158–65. Available from: http://dx.doi.org/10.1016/S2352-3018(20)30359-3

31. Yocum A, Simon EL. Thrombotic Thrombocytopenic Purpura after Ad26.COV2-S Vaccination. Am J Emerg Med [Internet]. 2021; Available from: https://www.sciencedirect.com/science/article/pii/S0735675721003764

